# Simulation-Guided Selection of a Bayesian Adaptive Phase II Design for a Nine-Arm Cilostazol-Albumin Trial in Aneurysmal Subarachnoid Hemorrhage

**DOI:** 10.64898/2026.06.18.26356019

**Authors:** Adnan I. Qureshi, Hassan Raza, Naima Alam, Jonathan Beall, Byron J. Gajewski, Renee L. Martin, Jose I. Suarez

**Affiliations:** Zeenat Qureshi Stroke Institute, University of Missouri, Columbia, Missouri, USA; Department of Neurology, University of Missouri, Columbia, Missouri, USA; Department of Biostatistics & Data Science, University of Kansas Medical Center, Kansas City, Kansas, USA; Department of Public Health Sciences, Medical University of South Carolina, Charleston, South Carolina, USA; Division of Neurosciences Critical Care, Departments of Anesthesiology and Critical Care Medicine, Neurology, and Neurosurgery, The Johns Hopkins University School of Medicine, Baltimore, Maryland, USA

**Keywords:** Bayesian adaptive design, response adaptive randomization, multi-arm Phase II trial, subarachnoid hemorrhage, simulation, dose-combination selection, dynamic linear model

## Abstract

**Background:** The Cilostazol Albumin Treatment in Subarachnoid Hemorrhage (CATS) trial evaluates eight active cilostazol-human albumin regimens plus control in patients with aneurysmal subarachnoid hemorrhage. We summarized the rationale for the primary statistical design, compared alternative Phase II methodologies, and evaluated reduced-arm sensitivity scenarios.

**Methods:** The binary primary endpoint is Common Data Elements-defined delayed cerebral ischemia within 14 days after randomization. The selected design is Bayesian adaptive, with a burn-in phase, response-adaptive randomization among active arms while maintaining fixed control allocation, four interim analyses, early stopping for expected success or futility, and a two-dimensional normal dynamic linear model. Primary operating characteristics were obtained from 1,000 virtual trials per scenario using Fixed and Adaptive Clinical Trial Simulator version 7.0.0. Exploratory simulations evaluated six-, four-, and two-active-arm configurations and simplified alternative designs.

**Results:** Compared with fixed equal allocation, the Bayesian adaptive design preserved an approximately 10% false-success probability under the global null while improving probability of success and efficiency in clinically relevant scenarios. Under the Realistic scenario, probability of success increased from 0.61 to 0.86, expected sample size decreased from 400 to 308, and expected duration decreased from 235 to 187 weeks. Under common thresholds, null probability of success was 0.098 for the full anchor and 0.073 for Reduced-6; Reduced-6 probabilities of success were 0.774 and 0.765 in the Realistic and Realistic2 scenarios. However, Reduced-6 omitted two monotherapy anchors and was less robust in Backwards2. In the comparator simulation, the selected design had probability of success of 0.858 and expected sample size of 308.3 under the Realistic scenario, compared with 0.624 to 0.845 and approximately 352 to 400 for simplified comparators.

**Conclusions:** For identifying the most promising cilostazol-human albumin regimen for Phase III rather than confirming efficacy, the Bayesian response-adaptive design with two-dimensional normal dynamic linear model borrowing is more efficient and better aligned than simplified comparators. The full nine-arm design remains preferable because it preserves the complete therapeutic discovery space and is more robust to misspecified or non-smooth response surfaces.

## Background

The Cilostazol Albumin Treatment in Subarachnoid Hemorrhage (CATS) trial is planned as a multicenter, randomized, double-blind, Bayesian adaptive Phase II trial in patients with aneurysmal subarachnoid hemorrhage (aSAH). aSAH remains a high-risk neurocritical illness in which delayed cerebral ischemia (DCI) is a clinically consequential complication, and American Heart Association/American Stroke Association (AHA/ASA) guidance emphasizes prevention, detection, and management of DCI [1–4]. The statistical challenge is typical of complex early-phase comparative effectiveness and dose-combination studies that require comparing multiple active regimens with a shared control, identifying the most promising regimen for Phase III evaluation, limiting unnecessary exposure to poorly performing regimens, accounting for multiplicity induced by multiple active-control comparisons, and preserving interpretability for clinical and regulatory decision-making. Adaptive designs are well suited to this setting when design adaptations, decision rules, simulation calibration, and reporting conventions are prespecified [5–7]. We summarize the design simulation results supporting the selected CATS statistical design and compare the selected approach with alternative Phase II methodologies, including fixed equal allocation, Simon two-stage screening [8], randomized Phase II selection [9–11], multi-arm multi-stage (MAMS) designs [12–16], factorial designs [17–19], Multiple Comparison Procedures and Modeling (MCP-Mod)/model-based dose-finding [20, 21], platform/master-protocol approaches [22–24], and Bayesian response-adaptive designs [25–31].

## Methods

### Clinical Design Problem

CATS evaluates nine randomized groups: one control arm and eight active cilostazol (CTZ)/25% human albumin (HA) regimens. The active regimens represent CTZ dose and HA duration components: CTZ 200 mg (CTZ200), CTZ 300 mg (CTZ300), HA for 1 day, HA for 7 days, and four CTZ/HA combinations (Table 1). This regimen set reflects prior clinical interest in CTZ for prevention of delayed ischemic complications after aSAH and pilot evidence supporting feasibility and safety evaluation of HA in aSAH [32–34].

**Table 1.** CATS randomized treatment groups.

| **Arm** | **Regimen description** | **Role in design** |
| --- | --- | --- |
| Control | Standard care / control regimen | Reference arm |
| CTZ200 | Cilostazol 200 mg for 14 days | Single-component active arm |
| CTZ300 | Cilostazol 300 mg for 14 days | Single-component active arm |
| HA1 | 25% human albumin 1.25 g/kg/day for 1 day | Single-component active arm |
| HA7 | 25% human albumin 1.25 g/kg/day for 7 days | Single-component active arm |
| CTZ200HA1 | CTZ 200 mg + HA for 1 day | Combination arm |
| CTZ200HA7 | CTZ 200 mg + HA for 7 days | Combination arm |
| CTZ300HA1 | CTZ 300 mg + HA for 1 day | Combination arm |
| CTZ300HA7 | CTZ 300 mg + HA for 7 days | Combination arm |

Abbreviations: CATS, Cilostazol Albumin Treatment in Subarachnoid Hemorrhage; CTZ, cilostazol; HA, 25% human albumin.

### Primary endpoint and Phase II estimand

The primary endpoint is National Institute of Neurological Disorders and Stroke Common Data Elements (NINDS CDE)-defined delayed cerebral ischemia (CDE-d-DCI) within 14 days post-randomization using central blinded interpretation of head computed tomography (CT) imaging. This endpoint choice is consistent with consensus efforts to standardize DCI-related outcome events and NINDS CDE for neurologic clinical research [1–3]. Lower event rates are better. The Phase II objective is to identify the most promising CTZ dose/HA duration regimen across the dose-combination grid for potential Phase III evaluation, rather than to confirm definitive efficacy of each active arm.

The final Go criterion requires a posterior probability greater than 0.97 that the best active arm has a lower event rate than control: Pr(*p_best_*< *p_control_* | data) > 0.97, where Pr denotes probability, *p_best_* denotes the active arm with the most favorable posterior profile, and *p_control_* denotes the control event rate. The early success criterion is more stringent, Pr(*p_best_*< *p_control_* | data) > 0.9925. These thresholds were selected through a trial-and-error calibration process to achieve an approximately 10% Type I error, as prespecified in the CATS Statistical Analysis Plan (SAP; version 1.0, 13 February 2025; unpublished). The main statistical design, including the two-dimensional smoothing model, is described in detail in a related manuscript by Alam et al., “Optimizing Combination Therapies Using a Bayesian Adaptive Design with a Two-dimensional NDLM” (manuscript under review). Early futility may be declared if the best-performing active arm has a posterior probability below 0.015 of achieving at least a 15-percentage-point reduction in the event rate relative to control.

### Selected Bayesian adaptive design

The selected design uses an initial burn-in period of 198 participants with block randomization in a 6:2:2:2:2:2:2:2:2 ratio across control and eight active arms (Table 2). After burn-in, response adaptive randomization (RAR) is applied among active arms while maintaining fixed allocation to control. The response-adaptive algorithm favors regimens with higher posterior probability of being the best active option while preventing continued allocation to arms with minimal probability of being best. Interim analyses are planned after approximately 198, 250, 300, and 350 participants, with a maximum sample size of 400. A two-dimensional normal dynamic linear model (2D NDLM) is used to model the response surface over the CTZ dose by HA duration grid. The model borrows information across neighboring active arms while modeling control separately. This approach is attractive because the eight active regimens are scientifically related rather than unrelated experimental products, but it requires sensitivity analysis because excessive smoothing can reduce robustness when the true response surface is irregular [30, 31, 35].

**Table 2.**
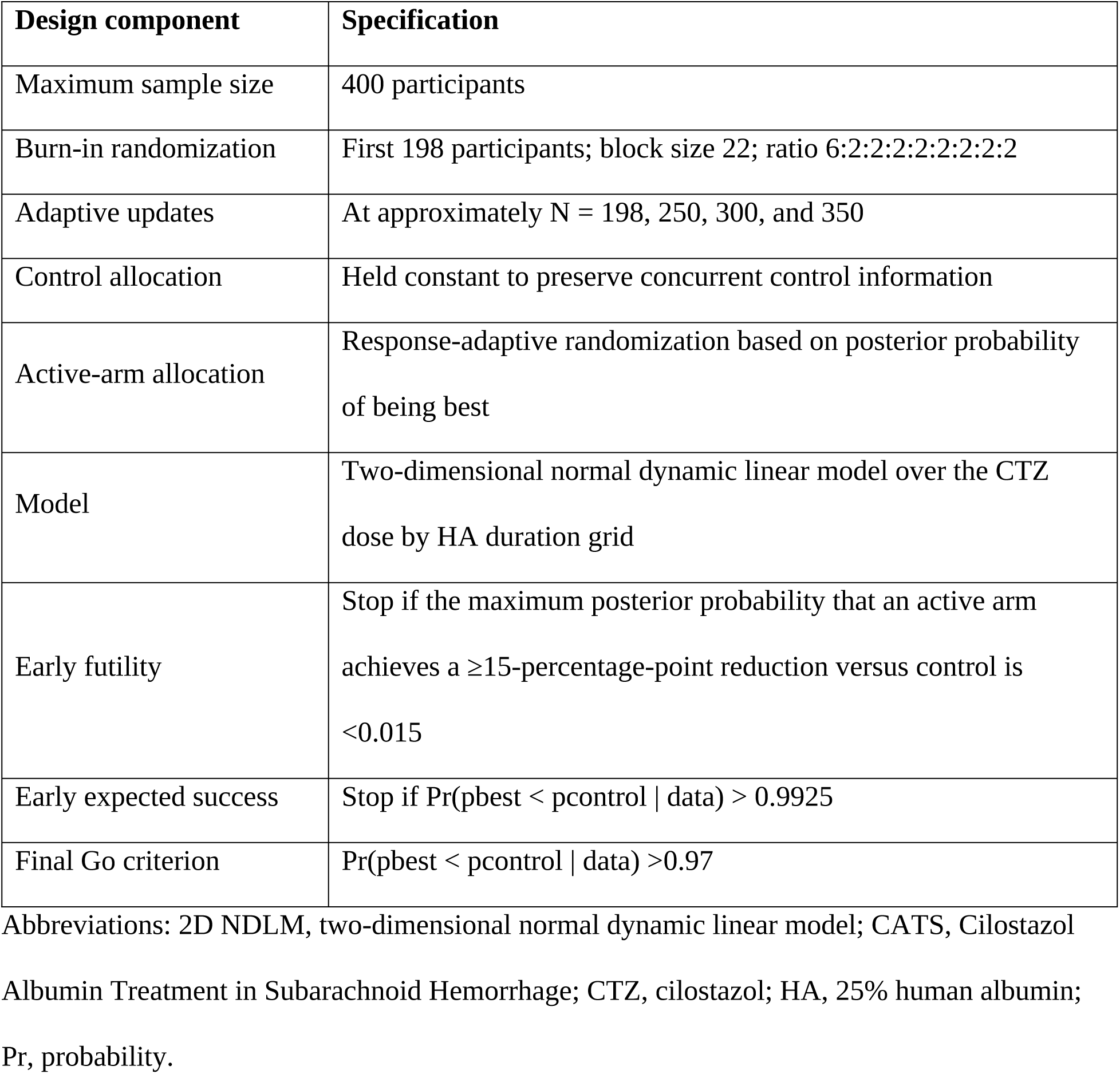
Key features of the selected Bayesian adaptive CATS design.

### Simulation framework

Operating characteristics were evaluated by virtual trial simulation using 1,000 simulated trials per scenario, generated under prespecified endpoint profiles, accrual assumptions, and dropout assumptions, as documented in the SAP. The primary simulations assumed accrual from a Poisson process with average accrual of approximately 1.92 participants per week and no dropouts. Fixed and Adaptive Clinical Trial Simulator (FACTS) version 7.0.0 was used for the adaptive trial simulations, and R was used to summarize output. Five high-level scenarios compared fixed and adaptive designs: null/no active benefit, all active arms beneficial, two realistic dose-combination response surfaces, and harmful active arms. The expanded operating-characteristic evaluation included nine virtual-subject response scenarios: All15, Backwards, Backwards2, Flat Drop, Harm10, Null, One Good One Bad, Realistic, and Realistic2.

### Reduced-arm sensitivity framework

An exploratory reduced-arm sensitivity analysis evaluated whether therapeutic arms could be removed while preserving the Phase II selection objective. Candidate configurations retained six, four, or two active arms under the same maximum sample size (Nmax) of 400. The analysis used the virtual-subject response profiles but did not rerun the original FACTS 2D NDLM implementation; instead, it used screening simulations with independent beta-binomial posterior approximations. Screening decision thresholds were calibrated in the full eight-active-arm model to approximate the SAP null operating characteristics and then applied unchanged to Reduced-6, Reduced-4, and Reduced-2. Thus, the reduced configurations were not separately recalibrated to the same achieved Type I error. These analyses describe the effect of arm removal under a common decision rule and are intended to identify promising or fragile configurations for formal FACTS/2D NDLM reruns, not to replace the prespecified SAP operating characteristics.

### Comparator Phase II methodologies

Comparator designs were selected to represent common early-phase approaches that could plausibly be considered for a nine-arm regimen-selection study. These included fixed equal-allocation multi-arm trials, Simon two-stage and parallel two-stage screening designs, randomized selection/pick-the-winner designs, frequentist group-sequential multi-arm designs, MAMS designs, factorial designs, MCP-Mod/model-based dose-finding approaches, platform or master-protocol designs, and Bayesian response-adaptive designs. The rationale, limitations, and key references for each comparator design are summarized in Supplementary Table 1.

### Monte Carlo design-comparator sensitivity simulation

We performed an additional Monte Carlo design-comparator sensitivity simulation. For each virtual participant, the primary endpoint was generated as a Bernoulli outcome with the scenario-specific CDE-d-DCI probability. The Bayesian adaptive RAR plus 2D NDLM operating characteristics were retained from the prespecified FACTS simulations. Comparator designs were implemented as transparent screening approximations: fixed final-only multi-arm screening, MAMS/drop-the-loser screening, parallel two-stage arm screening, and an independent-arm Bayesian RAR screen. The non-FACTS comparator designs were calibrated under the global null to approximately 10% false-success probability to avoid overstating benefit from relaxed decision thresholds. Metrics included probability of success, expected sample size, expected duration using the accrual assumption, futility stopping, and selection of a truly best regimen. These simulations are intended as a methodological sensitivity analysis and do not replace a definitive FACTS/2D NDLM rerun. The supplementary material provides the comparator-method details, Monte Carlo design-comparator sensitivity results, and complete Python code.

## Results

### Comparison with fixed equal allocation

The Bayesian adaptive design preserved the same approximate 0.10 one-sided false-success probability as the fixed design under the complete null scenario while materially improving power and efficiency under clinically relevant treatment-effect scenarios (Table 3 and Figure 1). Under the realistic scenario, probability of success increased from 0.61 with fixed equal allocation to 0.86 with the adaptive design; expected sample size decreased from 400 to 308; and expected duration decreased from 235 to 187 weeks. Under the all-active-beneficial scenario, probability of success increased from 0.78 to 0.94, expected sample size decreased to 268, and expected duration decreased to 166 weeks.

**Table 3.**
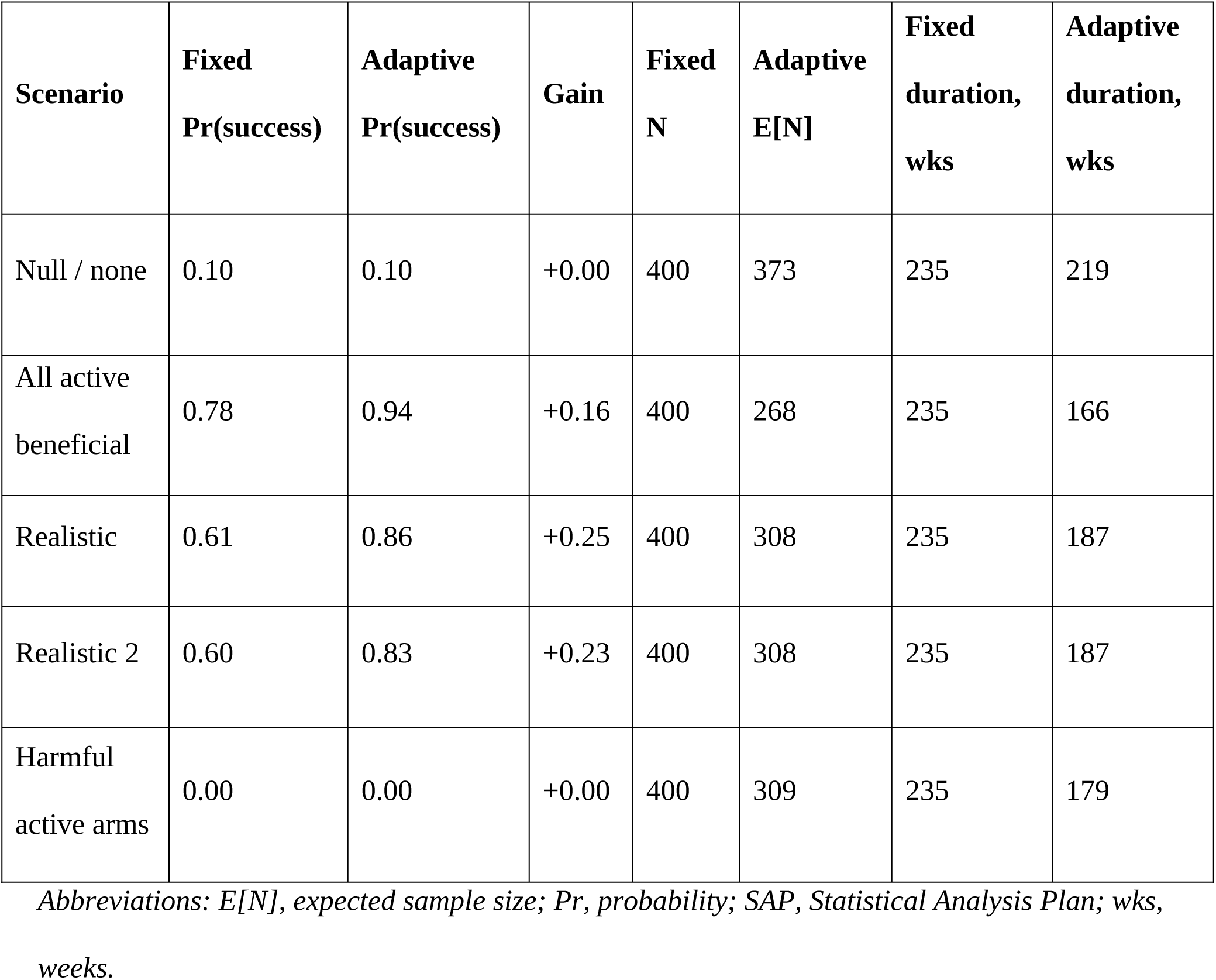
Fixed equal-allocation versus Bayesian adaptive design performance in five SAP scenarios.

**Figure 1.**
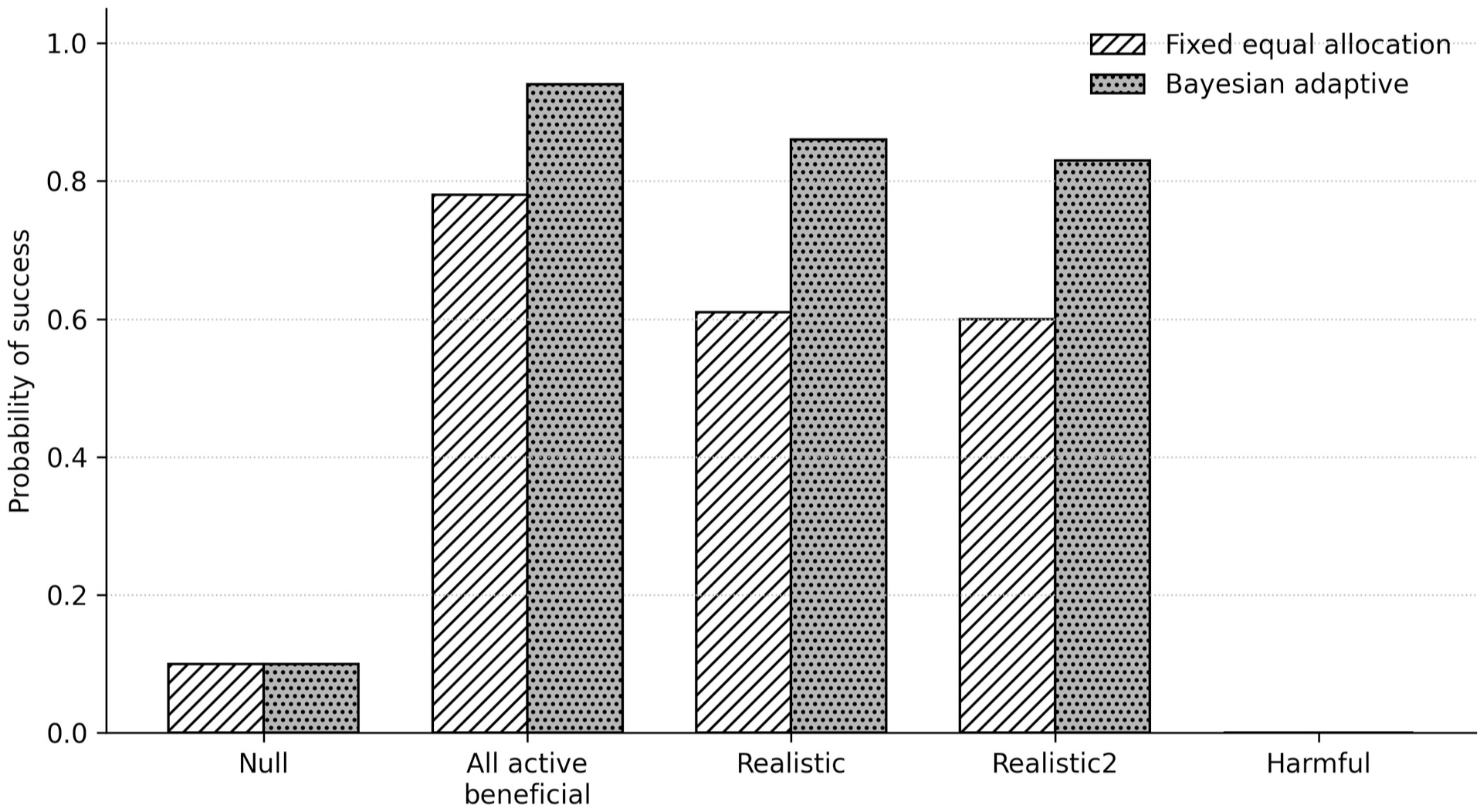
Success probability under fixed and adaptive designs. The Bayesian adaptive design increases probability of success under beneficial and realistic scenarios while maintaining approximately the same false-success probability under the global null. Harmful denotes the Harm10 scenario.

### Expanded adaptive-design operating characteristics

Across the nine expanded simulation scenarios, the adaptive design demonstrated high probability of success when one or more active regimens had clinically meaningful reductions in CDE-d-DCI, very low probability of success under harmful active-arm profiles, and expected sample sizes below the 400-participant maximum in all scenarios (Table 4). The Backwards scenario had a probability of success just below 0.80, suggesting sensitivity to irregular response surfaces and supporting prespecified sensitivity analyses with reduced borrowing.

**Table 4.**
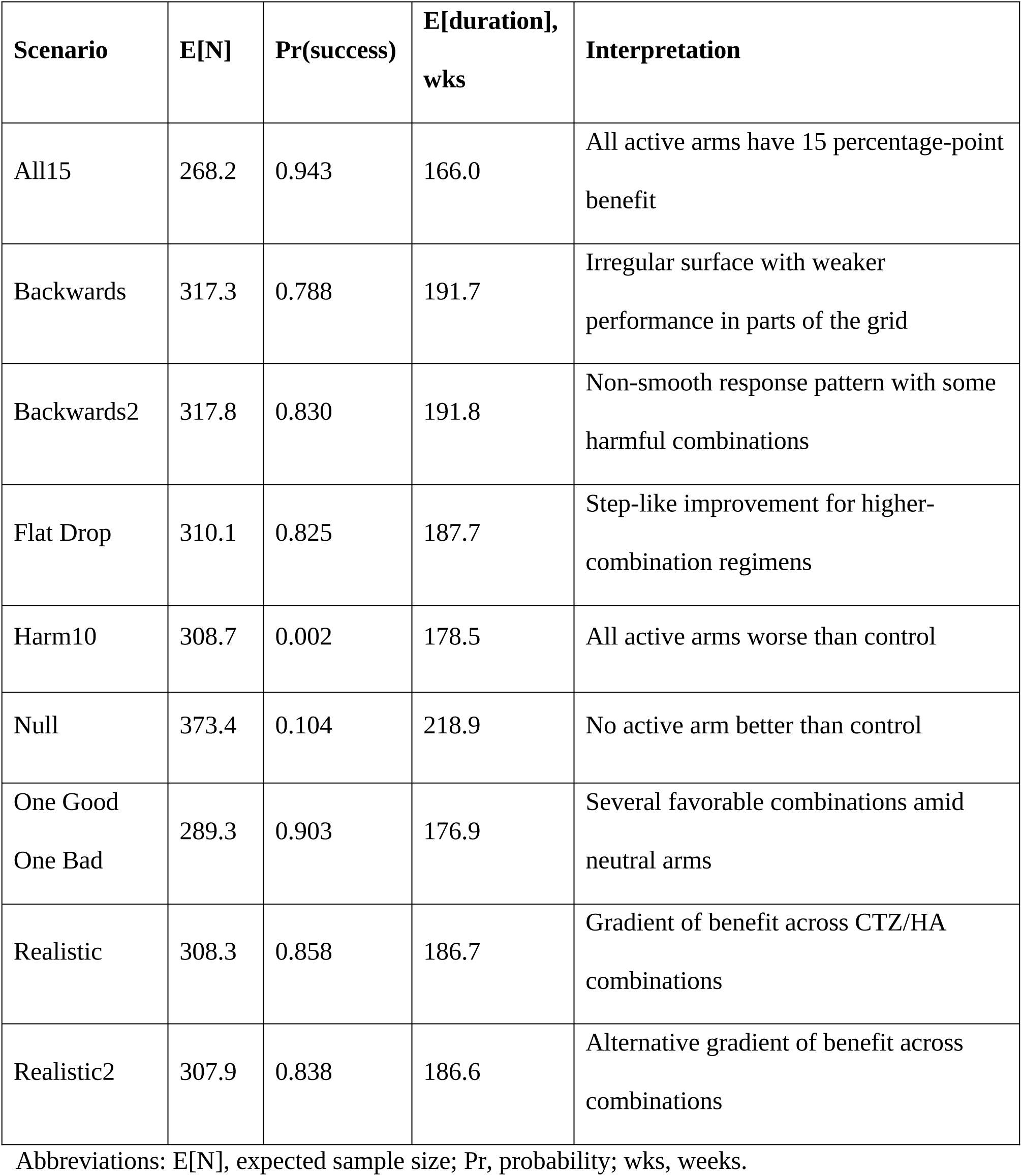
Expanded Bayesian adaptive operating characteristics from 1000 virtual trials per scenario.

| **Scenario** | **E[N]** | **Pr(success)** | **E[duration],**<br>**wks** | **Interpretation** |
| --- | --- | --- | --- | --- |
| All15 | 268.2 | 0.943 | 166.0 | All active arms have 15 percentage-point benefit |
| Backwards | 317.3 | 0.788 | 191.7 | Irregular surface with weaker performance in parts of the grid |
| Backwards2 | 317.8 | 0.830 | 191.8 | Non-smooth response pattern with some harmful combinations |
| Flat Drop | 310.1 | 0.825 | 187.7 | Step-like improvement for higher-combination regimens |
| Harm10 | 308.7 | 0.002 | 178.5 | All active arms worse than control |
| Null | 373.4 | 0.104 | 218.9 | No active arm better than control |
| One Good<br>One Bad | 289.3 | 0.903 | 176.9 | Several favorable combinations amid neutral arms |
| Realistic | 308.3 | 0.858 | 186.7 | Gradient of benefit across CTZ/HA combinations |
| Realistic2 | 307.9 | 0.838 | 186.6 | Alternative gradient of benefit across combinations |

Abbreviations: E[N], expected sample size; Pr, probability; wks, weeks.

### Reduced-arm sensitivity results

The reduced-arm sensitivity analysis assessed whether the original eight-active-arm therapeutic search space could be simplified while preserving the ability to identify a promising regimen. Because the original FACTS 2D NDLM file was not rerun and the reduced configurations were not separately recalibrated, these findings should be interpreted as common-threshold screening outputs rather than definitive configuration-specific operating characteristics. Under the same maximum sample size (Nmax) of 400 and common screening thresholds, the probability of success under the null scenario decreased as active arms were removed, largely because fewer arms could trigger a success decision (Table 5): 0.098 for the full eight-active-arm anchor, 0.073 for Reduced-6, 0.042 for Reduced-4, and 0.017 for Reduced-2. The probability of success under Harm10 was also very low across reduced configurations. These lower null values do not indicate superior calibration or inherent superiority. Moreover, omitted arms cannot be selected, contribute to component-effect interpretation, or inform the 2D response surface. Signal detection under clinically favorable profiles depended strongly on which arms were retained. Reduced-6, which retains selected monotherapy anchors and all four CTZ/HA combinations, had probability of success values of 0.774 and 0.765 in the Realistic and Realistic2 scenarios, respectively. Reduced-4 and Reduced-2 had similar or slightly higher values under smooth, combination-favorable assumptions, but they were fragile when the true response surface was not aligned with the retained arms. In the Backwards2 profile, probability of success was 0.668 for Reduced-6 and 0.743 for the full anchor; but only 0.011 and 0.004 for Reduced-4 and Reduced-2, respectively.

**Table 5.**
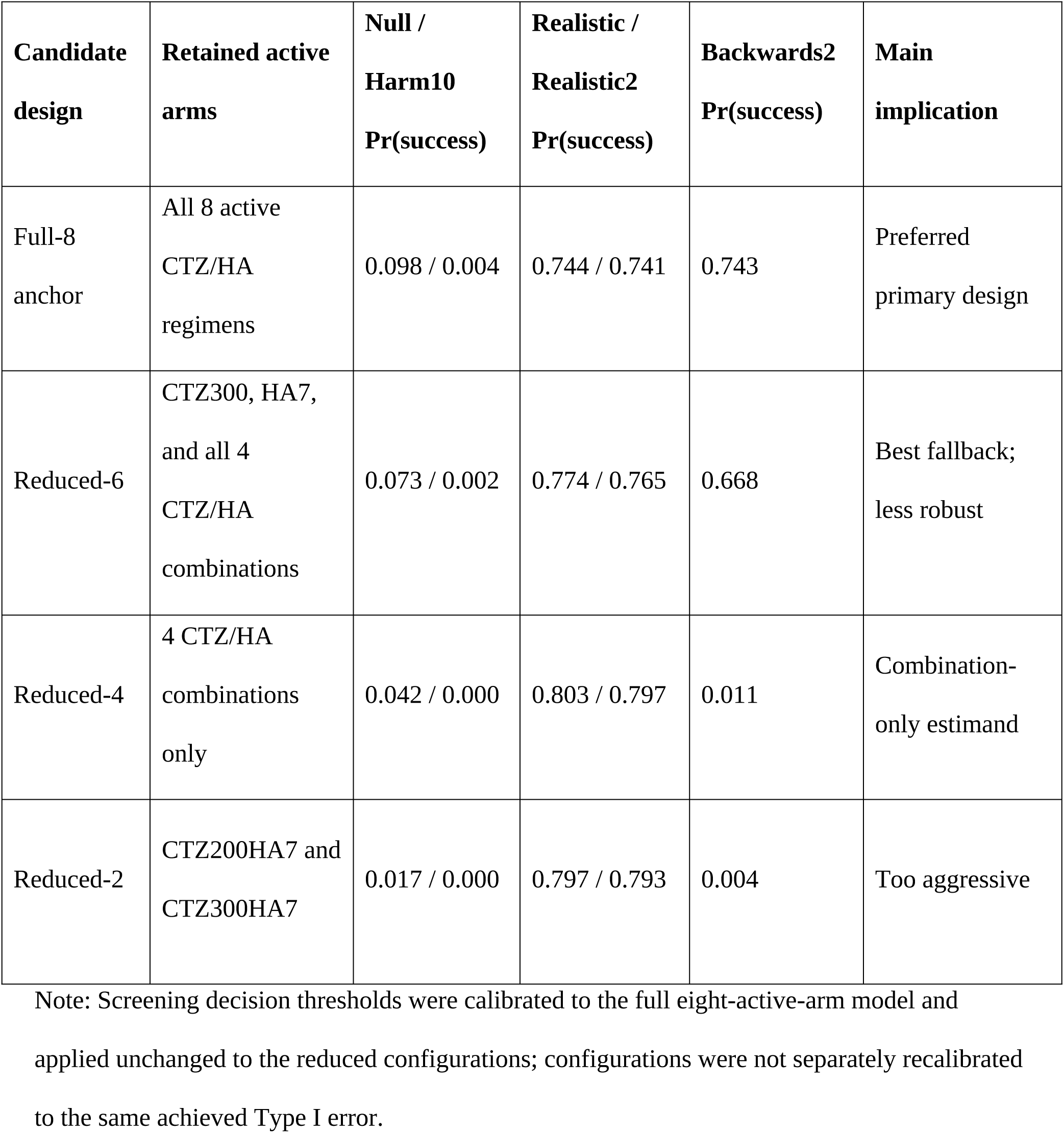
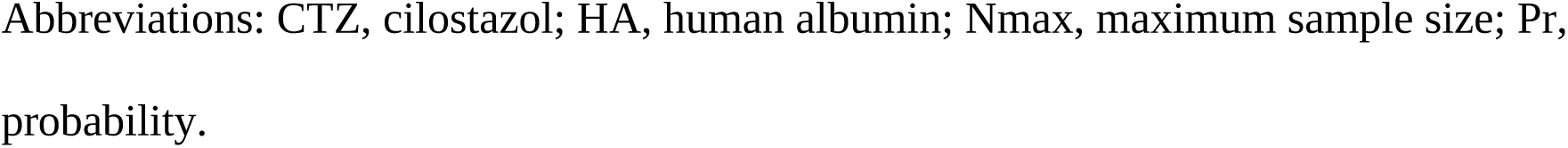
Reduced therapeutic-arm sensitivity results under Nmax = 400 using common screening decision thresholds calibrated to the full eight-active-arm anchor.

| **Candidate design** | **Retained active arms** | **Null / Harm10 Pr(success)** | **Realistic / Realistic2 Pr(success)** | **Backwards2 Pr(success)** | **Main implication** |
| --- | --- | --- | --- | --- | --- |
| Full-8 anchor | All 8 active CTZ/HA regimens | 0.098 / 0.004 | 0.744 / 0.741 | 0.743 | Preferred primary design |
| Reduced-6 | CTZ300, HA7, and all 4 CTZ/HA combinations | 0.073 / 0.002 | 0.774 / 0.765 | 0.668 | Best fallback; less robust |
| Reduced-4 | 4 CTZ/HA combinations only | 0.042 / 0.000 | 0.803 / 0.797 | 0.011 | Combination-only estimand |
| Reduced-2 | CTZ200HA7 and CTZ300HA7 | 0.017 / 0.000 | 0.797 / 0.793 | 0.004 | Too aggressive |

Note: Screening decision thresholds were calibrated to the full eight-active-arm model and applied unchanged to the reduced configurations; configurations were not separately recalibrated to the same achieved Type I error.
Abbreviations: CTZ, cilostazol; HA, human albumin; Nmax, maximum sample size; Pr, probability.

### Comparison of the full and reduced-arm designs

The sensitivity results support Reduced-6 as the best reduced-arm contingency, not as a replacement for the full nine-arm CATS design. Reduced-6 preserves all four CTZ/HA combination regimens and two monotherapy anchors while showing reasonable performance under smooth response profiles. However, its apparent advantages depend on the omitted arms being nonessential, meaning that certain CTZ doses or HA durations are unlikely to provide therapeutic benefit. The full nine-arm design is superior for the primary scientific objective for four related reasons. First, it preserves the complete therapeutic discovery space, including both single-component and combination regimens. Second, it retains CTZ200 and HA1 as lower-intensity component anchors, which are needed to interpret whether a favorable combination result is attributable to CTZ, HA, additive benefit, or interaction. Third, it supplies more complete local information for the 2D NDLM response surface, which is important when the true dose-combination relationship is not smooth or when a lower-intensity regimen is clinically preferable. Fourth, it provides more useful evidence for Phase III planning because it can support selection of an efficacious, safe, and practically deliverable regimen rather than only the best regimen among a narrowed set. In the Backwards2 sensitivity profile, Reduced-6 retained acceptable performance but was less robust than the full anchor (probability of success, 0.668 vs 0.743), while more restrictive reductions nearly eliminated the chance of success.

### Monte Carlo comparison with alternative designs

The Monte Carlo comparator simulation showed that the selected Bayesian adaptive CATS design provides the most favorable joint operating characteristics among the evaluated approaches (Supplementary Table 2). Under the Realistic scenario, the Bayesian adaptive design achieved a probability of success of 0.858 with an expected sample size of 308.3 and an expected duration of 186.7 weeks. By contrast, the fixed final-only screen had a probability of success of 0.624 with an expected sample size of 400, the MAMS/drop-the-loser screen had a probability of success of 0.845 but required an expected sample size of 399.1, and the parallel two-stage screen had a probability of success of 0.740 with an expected sample size of 399.7. The calibrated independent-arm Bayesian RAR screen reduced expected sample size to 351.5 but had a lower probability of success (0.749), illustrating that response-adaptive allocation alone is insufficient without the structured 2D NDLM borrowing and full simulation calibration used in the selected SAP design. Under the All15 scenario, in which all active regimens were beneficial, the SAP Bayesian adaptive design achieved the highest success probability (0.943) while reducing expected sample size to 268.2. MAMS/drop-the-loser screening was competitive for success probability (0.921) but did not materially reduce sample size when benefit was present (expected sample size of 399.7). Under the harmful-arm scenario, both the Bayesian design and MAMS produced very low false-success probabilities, but the Bayesian design had a shorter expected duration than the simulated MAMS and parallel two-stage comparators.

## Discussion

The simulation results support the Bayesian RAR design with 2D NDLM as the preferred primary statistical design. The design performed especially well in scenarios with smooth or partially smooth dose-combination response surfaces. In the Realistic and Realistic2 scenarios, the adaptive design increased probability of success by approximately 23 to 25 percentage points compared with a fixed equal-allocation design, while reducing expected sample size by approximately 92 participants and expected trial duration by approximately 48 weeks. These gains are important in aSAH, where eligible participants may be difficult to enroll and early identification of a promising regimen is valuable. The design also has ethical advantages. Fixed equal allocation intentionally maintains comparable sample sizes across arms even when accumulating evidence suggests that some regimens are unlikely to be optimal. In contrast, RAR shifts active-arm allocation toward better-performing regimens after the burn-in phase. By fixing control allocation rather than allowing the control arm to become underrepresented, the design mitigates a common limitation of RAR: excessive adaptation can weaken the precision of active-control comparisons.

The comparison with alternative designs supports the conclusion that the selected Bayesian design is superior because it combines calibrated decision thresholds, fixed concurrent control allocation, preferential allocation among active arms, early stopping, and model-based borrowing across the CTZ/HA grid. Fixed allocation preserves simplicity and interpretability but spends the full sample size regardless of early information and has substantially lower probability of success under realistic treatment effects. MAMS/drop-the-loser designs can protect against poor arms and were competitive for power, but the simulated MAMS approach did not provide meaningful sample-size or duration savings when active regimens were beneficial because allocation remained stage-fixed and early expected-success stopping was not intrinsic to the simplified design. Parallel two-stage screens reduced some exposure to harmful arms but had lower probability of success than the selected Bayesian design in realistic scenarios. The independent-arm RAR screen demonstrated that Bayesian updating and adaptive allocation alone are not sufficient; without borrowing over the CTZ dose by HA duration grid, stricter calibration is needed and power decreases. The selected design is therefore superior because it integrates four features simultaneously: concurrent control, calibrated posterior Go/No-Go rules, adaptive allocation, and structured 2D borrowing.

The reduced-arm sensitivity results have direct implications for protocol planning. The full nine-arm design remains superior to the Reduced-6 configuration as the primary scientific design, even though Reduced-6 is the preferred reduced-arm contingency. Under the common threshold calibrated to the full eight-active-arm anchor, Reduced-6 yielded a lower probability of success under the null scenario than the full anchor (0.073 vs 0.098), maintained very low probability of success in the Harm10 profile, preserved all four CTZ/HA combination regimens, and retained two monotherapy anchors. The lower null value reflects the smaller candidate search space and unequal achieved Type-I error; it does not indicate superior calibration or offset the information lost by removing arms. The price of simplification is loss of CTZ200 and HA1 monotherapy evidence and reduced ability to assess whether lower-intensity treatment could be effective, safer, or more practical for Phase III. The superiority of the full nine-arm design is particularly important for clinical and regulatory interpretation. A Phase III regimen recommendation should ideally be supported not only by a posterior probability that one retained arm is better than control, but also by evidence explaining why that regimen is preferred over plausible lower-intensity alternatives. Reduced-4 should be interpreted as a change in the clinical estimand rather than a simple efficiency improvement. A combination-only design may be attractive if the study team has already decided that only CTZ/HA combinations are plausible Phase III candidates; however, it would no longer answer the broader CATS question of whether any active CTZ or HA regimen is promising. Although Reduced-2 has the lowest null false-success probability under the common threshold, it provides too little protection against response-surface misspecification and would make the trial highly dependent on prior assumptions.

## Limitations

Several limitations should be recognized. First, the operating characteristics reported here depend on the simulation assumptions, including endpoint profiles, accrual model, and no-dropout assumption. Second, RAR is operationally and statistically complex; prior literature emphasizes that RAR can improve patient allocation in some settings but can also increase variability, bias, or logistical burden if poorly calibrated [25–29]. Third, the 2D NDLM assumes a degree of smoothness across related regimens. The Backwards scenario, with a probability of success of 0.788, indicates that performance can decrease when the true response surface is less smooth or partly counterintuitive [30, 31, 35]. Fourth, the SAP simulation results focus on probability of success, futility, sample size, trial duration, and allocation to best arms; future work should also quantify the probability of correctly selecting the truly best active regimen, posterior estimation bias, robustness to delayed endpoint availability, and safety-integrated decisions. Fifth, the reduced-arm sensitivity analysis is a common-threshold screening exercise rather than a definitive FACTS/2D NDLM rerun. Because the reduced configurations achieved different null false-success rates, favorable-scenario probability of success values should be interpreted descriptively rather than as direct power comparisons at a common Type I error. Accordingly, the reduced-arm results should be interpreted as screening evidence for prioritizing future configuration-specific recalibration and FACTS/2D NDLM reruns, not as final operating characteristics for protocol adoption.

## Conclusions

For a nine-arm Phase II CTZ/HA regimen-selection trial in aSAH, the Bayesian adaptive design with RAR, fixed control allocation, interim stopping, and 2D NDLM borrowing provides an efficient and decision-relevant framework for identifying the most promising regimen for Phase III evaluation. The full nine-arm design remains preferable to reduced-arm alternatives for the primary scientific question because it preserves the complete therapeutic discovery space, retains component and combination information, and provides stronger protection against misspecified or non-smooth response surfaces.

## Declarations

### Ethics approval and consent to participate

Not applicable. This methodological study analyzed trial-design simulations and did not involve human participants, human data, or human tissue.

### Consent for publication

Not applicable.

### Availability of data and materials

This study used simulation data and did not involve individual participant data. The scenario-level operating-characteristic data supporting the findings are presented in this article and the supplementary material. The supplementary material also contains the Python code used for the comparator simulations. The primary FACTS simulation summaries were derived from the CATS Statistical Analysis Plan.

### Authors’ contributions

AIQ conceived the study and supervised manuscript development. AIQ, and HR drafted the manuscript and prepared the tables, figure, and supplementary material. NA, JB, BJG, RLM, and JIS contributed to the statistical design and interpretation. All authors critically revised the manuscript, approved the final version, and agree to be accountable for the work.

## Acknowledgements

Not applicable.

## Supplementary material

Supplementary material: Comparator-methodology tables and reproducible Monte Carlo simulation code. Contains Supplementary Tables 1–3 and the complete Python code used for the design-comparator sensitivity simulation.

## List of abbreviations

2D NDLM: two-dimensional normal dynamic linear model
AHA/ASA: American Heart Association/American Stroke Association
aSAH: aneurysmal subarachnoid hemorrhage
CATS: Cilostazol Albumin Treatment in Subarachnoid Hemorrhage
CDE-d-DCI: Common Data Elements-defined delayed cerebral ischemia
CT: computed tomography
CTZ: cilostazol
DCI: delayed cerebral ischemia
FACTS: Fixed and Adaptive Clinical Trial Simulator
HA: human albumin
MAMS: multi-arm multi-stage
MCP-Mod: Multiple Comparison Procedures and Modeling
NINDS CDE: National Institute of Neurological Disorders and Stroke Common Data Elements
Nmax: maximum sample size
RAR: response-adaptive randomization
SAP: Statistical Analysis Plan.

## References

1. Vergouwen MDI, Vermeulen M, van Gijn J, Rinkel GJE, Wijdicks EF, Muizelaar JP, et al. Definition of delayed cerebral ischemia after aneurysmal subarachnoid hemorrhage as an outcome event in clinical trials and observational studies: proposal of a multidisciplinary research group. Stroke. 2010;41:2391–5. 10.1161/STROKEAHA.110.589275.

2. NINDS Common Data Elements | National Institute of Neurological Disorders and Stroke. https://www.ninds.nih.gov/current-research/scientific-resources/ninds-common-data-elements. Accessed 9 May 2026.

3. Frontera JA, Fernandez A, Schmidt JM, Claassen J, Wartenberg KE, Badjatia N, et al. Defining vasospasm after subarachnoid hemorrhage: what is the most clinically relevant definition? Stroke. 2009;40:1963–8. 10.1161/STROKEAHA.108.544700.

4. Hoh BL, Ko NU, Amin-Hanjani S, Chou SH-Y, Cruz-Flores S, Dangayach NS, et al. 2023 Guideline for the Management of Patients With Aneurysmal Subarachnoid Hemorrhage: A Guideline From the American Heart Association/American Stroke Association. Stroke. 2023;54:e314–70. 10.1161/STR.0000000000000436.

5. Adaptive Designs for Clinical Trials of Drugs and Biologics; Guidance for Industry; Availability. Federal Register. 2019. https://www.federalregister.gov/documents/2019/12/02/2019-25986/adaptive-designs-for-clinical-trials-of-drugs-and-biologics-guidance-for-industry-availability. Accessed 8 May 2026.

6. Dimairo M, Pallmann P, Wason J, Todd S, Jaki T, Julious SA, et al. The Adaptive designs CONSORT Extension (ACE) statement: a checklist with explanation and elaboration guideline for reporting randomised trials that use an adaptive design. BMJ. 2020;369:m115. 10.1136/bmj.m115.

7. Juszczak E, Altman DG, Hopewell S, Schulz K. Reporting of Multi-Arm Parallel-Group Randomized Trials: Extension of the CONSORT 2010 Statement. JAMA. 2019;321:1610–20. 10.1001/jama.2019.3087.

8. Simon R. Optimal two-stage designs for phase II clinical trials. Control Clin Trials. 1989;10:1–10. 10.1016/0197-2456(89)90015-9.

9. Simon R, Wittes RE, Ellenberg SS. Randomized phase II clinical trials. Cancer Treat Rep. 1985;69:1375–81.

10. Rubinstein LV, Korn EL, Freidlin B, Hunsberger S, Ivy SP, Smith MA. Design issues of randomized phase II trials and a proposal for phase II screening trials. J Clin Oncol. 2005;23:7199–206. 10.1200/JCO.2005.01.149.

11. Thall PF, Simon R, Ellenberg SS. Two-stage selection and testing designs for comparative clinical trials. Biometrika. 1988;75:303–10. 10.1093/biomet/75.2.303.

12. Royston P, Parmar MKB, Qian W. Novel designs for multi-arm clinical trials with survival outcomes with an application in ovarian cancer. Stat Med. 2003;22:2239–56. 10.1002/sim.1430.

13. Bratton DJ, Phillips PPJ, Parmar MKB. A multi-arm multi-stage clinical trial design for binary outcomes with application to tuberculosis. BMC Med Res Methodol. 2013;13:139. 10.1186/1471-2288-13-139.

14. Magirr D, Jaki T, Whitehead J. A generalized Dunnett test for multi-arm multi-stage clinical studies with treatment selection. Biometrika. 2012;99:494–501. 10.1093/biomet/ass002.

15. Jaki T, Magirr D. Considerations on covariates and endpoints in multi-arm multi-stage clinical trials selecting all promising treatments. Stat Med. 2013;32:1150–63. 10.1002/sim.5669.

16. Wason JMS, Trippa L. A comparison of Bayesian adaptive randomization and multi-stage designs for multi-arm clinical trials. Stat Med. 2014;33:2206–21. 10.1002/sim.6086.

17. Kahan BC, Hall SS, Beller EM, Birchenall M, Elbourne D, Juszczak E, et al. Consensus Statement for Protocols of Factorial Randomized Trials: Extension of the SPIRIT 2013 Statement. JAMA Netw Open. 2023;6:e2346121. 10.1001/jamanetworkopen.2023.46121.

18. Kahan BC, Hall SS, Beller EM, Birchenall M, Chan A-W, Elbourne D, et al. Reporting of Factorial Randomized Trials: Extension of the CONSORT 2010 Statement. JAMA. 2023;330:2106–14. 10.1001/jama.2023.19793.

19. Walter SD, Belo IJ. Design and analysis of factorial clinical trials: The impact of one treatment’s effectiveness on the statistical power and required sample size of the other. Stat Methods Med Res. 2023;32:1124–44. 10.1177/09622802231163332.

20. Bretz F, Pinheiro JC, Branson M. Combining multiple comparisons and modeling techniques in dose-response studies. Biometrics. 2005;61:738–48. 10.1111/j.1541-0420.2005.00344.x.

21. Pinheiro J, Bornkamp B, Glimm E, Bretz F. Model-based dose finding under model uncertainty using general parametric models. Stat Med. 2014;33:1646–61. 10.1002/sim.6052.

22. Research C for DE and. Master Protocols for Drug and Biological Product Development. 2023. https://www.fda.gov/regulatory-information/search-fda-guidance-documents/master-protocols-drug-and-biological-product-development. Accessed 9 May 2026.

23. Woodcock J, LaVange LM. Master Protocols to Study Multiple Therapies, Multiple Diseases, or Both. New England Journal of Medicine. 2017;377:62–70. 10.1056/NEJMra1510062.

24. Adaptive Platform Trials Coalition. Adaptive platform trials: definition, design, conduct and reporting considerations. Nat Rev Drug Discov. 2019;18:797–807. 10.1038/s41573-019-0034-3.

25. Thall PF, Wathen JK. Practical Bayesian adaptive randomisation in clinical trials. Eur J Cancer. 2007;43:859–66. 10.1016/j.ejca.2007.01.006.

26. Wathen JK, Thall PF. A simulation study of outcome adaptive randomization in multi-arm clinical trials. Clin Trials. 2017;14:432–40. 10.1177/1740774517692302.

27. Robertson DS, Lee KM, López-Kolkovska BC, Villar SS. Response-adaptive randomization in clinical trials: from myths to practical considerations. Stat Sci. 2023;38:185–208. 10.1214/22-STS865.

28. Proschan M, Evans S. Resist the Temptation of Response-Adaptive Randomization. Clin Infect Dis. 2020;71:3002–4. 10.1093/cid/ciaa334.

29. Villar SS, Robertson DS, Rosenberger WF. The Temptation of Overgeneralizing Response-adaptive Randomization. Clin Infect Dis. 2021;73:e842. 10.1093/cid/ciaa1027.

30. Berry DA, Müller P, Grieve AP, Smith M, Parke T, Blazek R, et al. Adaptive Bayesian Designs for Dose-Ranging Drug Trials. In: Gatsonis C, Kass RE, Carlin B, Carriquiry A, Gelman A, Verdinelli I, et al., editors. Case Studies in Bayesian Statistics. New York, NY: Springer; 2002. p. 99–181. 10.1007/978-1-4613-0035-9_2.

31. West M, Harrison J. Bayesian Forecasting and Dynamic Models. Springer; 1989.

32. Liu J, He J, Chen X, Feng Y, Wang C, Awil MA, et al. Cilostazol for Aneurysmal Subarachnoid Hemorrhage: An Updated Systematic Review and Meta-Analysis. Cerebrovasc Dis. 2022;51:138–48. 10.1159/000518731.

33. Saber H, Desai A, Palla M, Mohamed W, Seraji-Bozorgzad N, Ibrahim M. Efficacy of Cilostazol in Prevention of Delayed Cerebral Ischemia after Aneurysmal Subarachnoid Hemorrhage: A Meta-Analysis. J Stroke Cerebrovasc Dis. 2018;27:2979–85. 10.1016/j.jstrokecerebrovasdis.2018.06.027.

34. Suarez JI, Martin RH, Calvillo E, Dillon C, Bershad EM, Macdonald RL, et al. The Albumin in Subarachnoid Hemorrhage (ALISAH) multicenter pilot clinical trial: safety and neurologic outcomes. Stroke. 2012;43:683–90. 10.1161/STROKEAHA.111.633958.

35. Berry SM, Carlin BP, Lee JJ, Muller P. Bayesian Adaptive Methods for Clinical Trials. Boca Raton: CRC Press; 2010. 10.1201/EBK1439825488.

