## Supplementary for "Simulation-Guided Selection of a Bayesian Adaptive Phase II Design for a Nine-Arm Cilostazol-Albumin Trial in Aneurysmal Subarachnoid Hemorrhage"

#### Comparator-methodology tables and reproducible Monte Carlo simulation code

### Supplementary Appendix 1. Comparator-methodology and Monte Carlo sensitivity tables

This appendix contains the comparator-design table and Monte Carlo design-comparator sensitivity results cited in the main manuscript. The Monte Carlo comparator simulation evaluates simplified screening approximations and should not replace definitive FACTS/2D NDLM reruns.

##### Supplementary Table 1. Comparison of selected Bayesian adaptive CATS design with alternative Phase II methodologies.

| Methodology | Strengths | Limitations for CATS | Overall fit for CATS | Key refs |
|---|---|---|---|---|
| Fixed equal-allocation multi-arm randomized Phase II | Simple, transparent, familiar; concurrent control; unbiased allocation. | Enrolls many participants to inferior arms; no early stopping; no borrowing across structured regimen grid; lower power and longer expected duration in CATS simulations. | Acceptable but inefficient. Serves as benchmark, not preferred primary design. | [7,16] |
| Simon two-stage single-arm design | Efficient screening for one experimental regimen when historical control is reliable; minimizes expected sample size under poor activity. | Not designed for eight active regimens plus concurrent control; does not address active-arm selection, shared control, or multi-arm multiplicity; vulnerable to historical-control bias. | Poor fit. Could be useful for pre-CATS single-regimen screening only. | [8] |
| Multiple parallel two-stage designs | Could screen each regimen for lack of activity; simple within each regimen. | Inefficient use of control information; complex multiplicity; no adaptive prioritization; inflated resource burden for eight active regimens. | Inferior to a unified multi-arm adaptive design. | [8,10,11] |
| Randomized selection / pick-the-winner | Directly aligned with regimen selection; may be simpler than fully adaptive Bayesian design. | Often emphasizes ranking rather than calibrated evidence versus control; may not include early stopping, posterior Go/No-Go thresholds, or structured borrowing. | Partially aligned but less complete for CATS decision-making. | [9-11] |
| Frequentist group-sequential multi-arm design | Formal repeated testing framework; possible early stopping; strong control can be specified. | Less flexible for response-adaptive allocation and borrowing over a two-dimensional dose-combination grid; can be operationally complex for best-arm selection. | Viable alternative, but less efficient for CATS design goals unless augmented substantially. | [12-15] |
| MAMS / drop-the-loser design | Efficient evaluation of multiple arms against a common control; can drop poorly performing arms; strong tradition in multi-arm trials. | Typically allocates by fixed ratios within stages and does not preferentially assign more participants to better arms; borrowing across CTZ/HA grid is not intrinsic. | Strong comparator. Less aligned than Bayesian adaptive RAR + 2D NDLM for adaptive allocation and structured borrowing. | [12-16] |
| Factorial design | Can estimate main effects and interactions of CTZ and HA components; efficient when interaction assumptions are credible. | CATS has a regimen-level selection objective; CTZ doses and HA durations create a structured grid with plausible interactions; equal factorial allocation does not prioritize best regimens. | Useful as a secondary conceptual framework, but not ideal as primary Go/No-Go design. | [17-19] |
| MCP-Mod or model-based dose-response design | Efficient for ordered dose-response signal detection and dose estimation under model uncertainty. | Less natural for a binary outcome over two treatment components with combination regimens and regimen-selection objective; would require substantial adaptation. | Potential sensitivity tool, not preferred primary design. | [20,21] |
| Platform / master protocol design | Efficient for long-running disease programs with arms added/dropped over time and shared infrastructure. | CATS is a fixed nine-arm Phase II regimen-selection trial; a full platform structure adds governance and regulatory complexity without clear incremental benefit. | Could be considered for a future broader aSAH therapeutic program, not necessary for current CATS scope. | [22-24] |
| Bayesian adaptive RAR + 2D NDLM | Higher probability of success in realistic scenarios; lower expected sample size and duration; fixed control allocation; early success/futility; preferential active-arm allocation; structured borrowing across CTZ/HA grid. | Requires simulation-based calibration, unblinded statistical monitoring, careful Data and Safety Monitoring Board procedures, and sensitivity analyses for irregular response surfaces. | Best fit for the CATS Phase II selection objective. | [5-7,25-31,35] |

Abbreviations: 2D NDLM, two-dimensional normal dynamic linear model; CATS, Cilostazol Albumin Treatment in Subarachnoid Hemorrhage; CTZ, cilostazol; HA, human albumin; MAMS, multi-arm multi-stage; MCP-Mod, Multiple Comparison Procedures and Modeling; RAR, response-adaptive randomization.

**Supplementary Table 2. Monte Carlo comparison of selected Bayesian adaptive design with alternative Phase II screening designs.**

| Design | Null Pr(success) | All15 Pr(success)/E[N] | Realistic Pr(success)/E[N] | Realistic duration, wk | Harm10 Pr(success)/E[N] | Main implication |
|---|---|---|---|---|---|---|
| Bayesian adaptive RAR + 2D NDLM (SAP FACTS) | 0.104 | 0.943 / 268.2 | 0.858 / 308.3 | 186.7 | 0.002 / 308.7 | Best balance of power, sample size, and duration. |
| Fixed equal allocation (SAP) | 0.100 | 0.780 / 400.0 | 0.610 / 400.0 | 235.0 | 0.000 / 400.0 | Transparent benchmark but inefficient. |
| Fixed final-only Bayesian screen | 0.091 | 0.750 / 400.0 | 0.624 / 400.0 | 235.0 | 0.008 / 400.0 | Similar to fixed benchmark; no adaptive efficiency. |
| MAMS/drop-the-loser screen | 0.104 | 0.921 / 399.7 | 0.845 / 399.1 | 234.5 | 0.002 / 298.9 | Competitive power but little sample saving when benefit is present. |
| Parallel two-stage arm screens | 0.098 | 0.860 / 399.9 | 0.740 / 399.7 | 234.9 | 0.003 / 336.2 | Less powerful and slower than selected Bayesian design. |
| Bayesian independent-arm RAR screen | 0.095 | 0.882 / 323.3 | 0.749 / 351.5 | 209.8 | 0.002 / 359.3 | Shows that RAR without 2D borrowing is not enough. |

Note: FACTS values are from the prespecified CATS simulations; simplified comparator values were generated from 3000 Monte Carlo trials per scenario after null calibration. Abbreviations: 2D NDLM, two-dimensional normal dynamic linear model; CATS, Cilostazol Albumin Treatment in Subarachnoid Hemorrhage; E[N], expected sample size; FACTS, Fixed and Adaptive Clinical Trial Simulator; MAMS, multi-arm multi-stage; Pr, probability; RAR, response-adaptive randomization; SAP, Statistical Analysis Plan; wk, week.

### Supplementary Appendix 2. Reproducible Monte Carlo design-comparator Python code

Purpose. This appendix provides the Python code used to perform a Monte Carlo design-comparator sensitivity simulation for the CATS Phase II design manuscript. The script compares simplified alternative Phase II screening approaches against the CATS Bayesian adaptive response-adaptive randomization plus 2D NDLM design results from the SAP/FACTS simulation framework.

Important interpretation note. The selected CATS Bayesian adaptive RAR + 2D NDLM operating characteristics are entered as SAP/FACTS constants. The alternative comparator designs are transparent beta-binomial Monte Carlo approximations calibrated under the global null to an approximate 10% false-success rate. This script is intended for manuscript sensitivity analysis, not as a replacement for the full FACTS/2D NDLM trial simulator.

Software requirements. Python 3.x with numpy, pandas, and scipy installed.

Script metadata are summarized in Supplementary Table 3.

**Supplementary Table 3. Python script metadata.**

| **Source script** | **CATS_monte_carlo_design_comparison.py** |
|---|---|
| Number of code lines | 296 |
| Primary output | /mnt/data/cats_monte_carlo_design_comparison.csv |
| Main command | python CATS_monte_carlo_design_comparison.py |
| Randomness | Seeded Monte Carlo simulations for reproducibility |

Abbreviations: 2D NDLM, two-dimensional normal dynamic linear model; CATS, Cilostazol Albumin Treatment in Subarachnoid Hemorrhage; FACTS, Fixed and Adaptive Clinical Trial Simulator; MAMS, multi-arm multi-stage; RAR, response-adaptive randomization; SAP, Statistical Analysis Plan.

**Designs represented in the script:**

- CATS Bayesian adaptive RAR + 2D NDLM (SAP/FACTS results entered as constants)
- CATS fixed equal allocation (SAP benchmark results entered as constants)
- Fixed final-only Bayesian screen
- MAMS/drop-the-loser screen
- Parallel two-stage arm screens
- Bayesian independent-arm RAR screen

#### Complete Python code listing

```
import numpy as np
import pandas as pd
import hashlib
from scipy.stats import norm

ARM_NAMES = ["Control", "CTZ200", "CTZ300", "HA1", "HA7", "CTZ200HA1", "CTZ200HA7", "CTZ300HA1", "CTZ300HA7"]
SCENARIOS = {
    "All15":       [0.31,0.16,0.16,0.16,0.16,0.16,0.16,0.16,0.16],
    "Backwards":   [0.31,0.20,0.23,0.20,0.23,0.16,0.18,0.27,0.18],
    "Backwards2":  [0.31,0.17,0.16,0.17,0.16,0.34,0.34,0.34,0.34],
    "Flat Drop":   [0.31,0.23,0.23,0.23,0.23,0.18,0.18,0.17,0.16],
    "Harm10":      [0.31,0.41,0.41,0.41,0.41,0.41,0.41,0.41,0.41],
    "Null":        [0.31,0.31,0.31,0.31,0.31,0.31,0.31,0.31,0.31],
    "One Good One Bad":[0.31,0.31,0.31,0.17,0.16,0.17,0.16,0.17,0.16],
    "Realistic":   [0.31,0.27,0.25,0.23,0.20,0.18,0.17,0.17,0.16],
    "Realistic2":  [0.31,0.23,0.20,0.27,0.25,0.17,0.16,0.18,0.17],
}
SAP_FACTS = {
    "All15":       {"E_N":268.2,"Pr_success":0.943,"E_duration":166.0},
    "Backwards":   {"E_N":317.3,"Pr_success":0.788,"E_duration":191.7},
    "Backwards2":  {"E_N":317.8,"Pr_success":0.830,"E_duration":191.8},
    "Flat Drop":   {"E_N":310.1,"Pr_success":0.825,"E_duration":187.7},
    "Harm10":      {"E_N":308.7,"Pr_success":0.002,"E_duration":178.5},
    "Null":        {"E_N":373.4,"Pr_success":0.104,"E_duration":218.9},
```

```
    "One Good One Bad":{"E_N":289.3,"Pr_success":0.903,"E_duration":176.9},
    "Realistic":   {"E_N":308.3,"Pr_success":0.858,"E_duration":186.7},
    "Realistic2":  {"E_N":307.9,"Pr_success":0.838,"E_duration":186.6},
}
SAP_FIXED = {
    "Null":      {"E_N":400.0,"Pr_success":0.10,"E_duration":235.0},
    "All15":     {"E_N":400.0,"Pr_success":0.78,"E_duration":235.0},
    "Realistic": {"E_N":400.0,"Pr_success":0.61,"E_duration":235.0},
    "Realistic2":{"E_N":400.0,"Pr_success":0.60,"E_duration":235.0},
    "Harm10":    {"E_N":400.0,"Pr_success":0.00,"E_duration":235.0},
}

def beta_mean_var(y, n):
    a = y + 1.0
    b = n - y + 1.0
    mean = a / (a+b)
    var = (a*b)/(((a+b)**2)*(a+b+1.0))
    return mean, var

def pp_better(y_active, n_active, y_ctrl, n_ctrl):
    ma,va=beta_mean_var(y_active,n_active); mc,vc=beta_mean_var(y_ctrl,n_ctrl)
    return norm.cdf((mc-ma)/np.sqrt(va+vc))

def pp_abs_reduction(y_active, n_active, y_ctrl, n_ctrl, delta=0.15):
    ma,va=beta_mean_var(y_active,n_active); mc,vc=beta_mean_var(y_ctrl,n_ctrl)
    return norm.cdf(((mc-ma)-delta)/np.sqrt(va+vc))

def eval_results(p, success, N, selected, pct_top4=None, futility=None):
    p=np.array(p)
    true_best=np.where(p[1:]==np.min(p[1:]))[0]+1
    correct=(np.isin(selected,true_best) & success)
    dur=N/1.92 + 26.6666666667
    out={
        "Pr_success":float(np.mean(success)),
        "E_N":float(np.mean(N)),
        "E_duration":float(np.mean(dur)),
        "Pr_select_true_best":float(np.mean(correct)),
        "Pr_futility":float(np.mean(futility)) if futility is not None else 0.0,
        "Pct_top4_alloc":float(np.nanmean(pct_top4)) if pct_top4 is not None else np.nan,
    }
    return out

def sim_fixed_final(p, nsim=10000, threshold=0.97, seed=1):
    rng=np.random.default_rng(seed); p=np.array(p)
    counts=np.array([45,45,45,44,44,44,44,44,43])
    n=np.broadcast_to(counts,(nsim,9)).copy()
    y=rng.binomial(n,p)
    pp=pp_better(y[:,1:],n[:,1:],y[:,[0]],n[:,[0]])
    success=pp.max(axis=1)>threshold
    selected=pp.argmax(axis=1)+1
    N=np.full(nsim,400.0)
    top4=np.argsort(p[1:])[:4]+1
    pct_top4=np.full(nsim,counts[top4].sum()/400.0)
    return eval_results(p,success,N,selected,pct_top4=pct_top4)

def sim_mams(p, nsim=10000, threshold=0.97, futility_pp=0.55, seed=2):
    rng=np.random.default_rng(seed); p=np.array(p)
    y=np.zeros((nsim,9),dtype=int); n=np.zeros((nsim,9),dtype=int)
    n1=np.array([54]+[18]*8)
    y += rng.binomial(n1,p,size=(nsim,9)); n += n1
    pp=pp_better(y[:,1:],n[:,1:],y[:,[0]],n[:,[0]])
    fut1=pp.max(axis=1)<futility_pp
    active4=np.argsort(pp,axis=1)[:,-4:]+1  # top 4, unsorted
    # Stage2 add 102: 22 control, 20 or 21 to selected arms deterministic pattern [21,21,20,20]
    alive=~fut1
    n[alive,0]+=22; y[alive,0]+=rng.binomial(22,p[0],size=alive.sum())
    add4=np.array([20,20,20,20])
    rows=np.where(alive)[0]
    for j in range(4):
        arms=active4[rows,j]
        n[rows,arms]+=add4[j]
        y[rows,arms]+=rng.binomial(add4[j],p[arms])
    # Stage2 decision
    pp2_all=np.full((nsim,4),np.nan)
    if len(rows)>0:
        pp2_all[rows]=pp_better(y[rows[:,None],active4[rows]], n[rows[:,None],active4[rows]], y[rows,[0]][:,None], n[rows,[0]][:,None])
```

```
    fut2=np.zeros(nsim,dtype=bool)
    fut2[alive]=np.nanmax(pp2_all[alive],axis=1)<futility_pp
    active2=np.zeros((nsim,2),dtype=int)
    idx_sort=np.argsort(pp2_all,axis=1)[:,-2:]
    for i in rows:
        active2[i]=active4[i,idx_sort[i]]
    alive2=alive & ~fut2
    # Stage3 add 100: 34 control, 33 and 33 to selected arms
    rows2=np.where(alive2)[0]
    n[rows2,0]+=34; y[rows2,0]+=rng.binomial(34,p[0],size=len(rows2))
    for j in range(2):
        arms=active2[rows2,j]
        n[rows2,arms]+=33
        y[rows2,arms]+=rng.binomial(33,p[arms])
    # Final
    pp_final=np.zeros((nsim,2))
    if len(rows2)>0:
        pp_final[rows2]=pp_better(y[rows2[:,None],active2[rows2]], n[rows2[:,None],active2[rows2]], y[rows2,[0]][:,None], n[rows2,[0]][:,None])
    success=np.zeros(nsim,dtype=bool); success[rows2]=np.nanmax(pp_final[rows2],axis=1)>threshold
    selected=np.ones(nsim,dtype=int)
    # selected for stopped cases = top at stop; for completed cases = max final
    selected[fut1]=pp[fut1].argmax(axis=1)+1
    rows_fut2=np.where(fut2 & alive)[0]
    if len(rows_fut2)>0:
        selected[rows_fut2]=active4[rows_fut2,np.nanargmax(pp2_all[rows_fut2],axis=1)]
    if len(rows2)>0:
        selected[rows2]=active2[rows2,pp_final[rows2].argmax(axis=1)]
    N=n.sum(axis=1).astype(float)
    top4=np.argsort(p[1:])[:4]+1
    pct_top4=n[:,top4].sum(axis=1)/N
    futility=fut1|fut2
    return eval_results(p,success,N,selected,pct_top4=pct_top4,futility=futility)

def sim_parallel_two_stage(p, nsim=10000, threshold=0.97, keep_pp=0.50, futility_pp=0.45, seed=3):
    rng=np.random.default_rng(seed); p=np.array(p)
    y=np.zeros((nsim,9),dtype=int); n=np.zeros((nsim,9),dtype=int)
    n1=np.array([54]+[18]*8)
    y += rng.binomial(n1,p,size=(nsim,9)); n += n1
    pp=pp_better(y[:,1:],n[:,1:],y[:,[0]],n[:,[0]])
    keep=pp>=keep_pp
    fut=(keep.sum(axis=1)==0) | (pp.max(axis=1)<futility_pp)
    selected=pp.argmax(axis=1)+1
    alive=~fut
    # Continue to N=400. Allocate remaining equally among control and kept arms.
    rows=np.where(alive)[0]
    for i in rows:
        k=int(keep[i].sum())
        add_total=400-int(n[i].sum())
        # equal deterministic counts among control+k active
        base=add_total//(k+1); rem=add_total%(k+1)
        n0=base+(1 if rem>0 else 0)
        n[i,0]+=n0; y[i,0]+=rng.binomial(n0,p[0])
        active=np.where(keep[i])[0]+1
        for j,arm in enumerate(active):
            add=base+(1 if (j+1)<rem else 0)
            n[i,arm]+=add; y[i,arm]+=rng.binomial(add,p[arm])
        pp_active=pp_better(y[i,active],n[i,active],y[i,0],n[i,0])
        selected[i]=active[int(np.argmax(pp_active))]
    # final pp for selected arm only
    pp_sel=np.zeros(nsim)
    rows=np.where(alive)[0]
    if len(rows)>0:
        pp_sel[rows]=pp_better(y[rows,selected[rows]],n[rows,selected[rows]],y[rows,0],n[rows,0])
    success=np.zeros(nsim,dtype=bool); success[rows]=pp_sel[rows]>threshold
    N=n.sum(axis=1).astype(float)
    top4=np.argsort(p[1:])[:4]+1
    pct_top4=np.where(N>198,n[:,top4].sum(axis=1)/N,np.nan)
    return eval_results(p,success,N,selected,pct_top4=pct_top4,futility=fut)

def allocate_counts(weights, n_add):
    # weights shape (nsim,8), row-normalized. return integer counts sum n_add.
    raw=weights*n_add
    base=np.floor(raw).astype(int)
    rem=n_add-base.sum(axis=1)
    frac=raw-base
    # add remaining to highest fractional weights
```

```
    order=np.argsort(frac,axis=1)[:,::-1]
    for r in range(weights.shape[0]):
        if rem[r]>0:
            base[r,order[r,:rem[r]]]+=1
        elif rem[r]<0:
            # shouldn't occur; remove from smallest fractional positive counts
            candidates=np.where(base[r]>0)[0]
            for a in candidates[:(-rem[r])]:
                base[r,a]-=1
    return base

def sim_bayes_ind_rar(p, nsim=10000, threshold=0.97, early_success=0.9925, early_futility=0.015, seed=4):
    rng=np.random.default_rng(seed); p=np.array(p)
    y=np.zeros((nsim,9),dtype=int); n=np.zeros((nsim,9),dtype=int)
    n1=np.array([54]+[18]*8)
    y += rng.binomial(n1,p,size=(nsim,9)); n += n1
    active=np.ones(nsim,dtype=bool)
    success=np.zeros(nsim,dtype=bool); fut=np.zeros(nsim,dtype=bool); selected=np.ones(nsim,dtype=int)
    blocks=[(250,14),(300,14),(350,14),(400,14)]  # target, control add approx 6/22
    for b,(target,n_ctrl_add) in enumerate(blocks):
        rows=np.where(active)[0]
        if len(rows)==0: break
        pp=pp_better(y[rows,1:],n[rows,1:],y[rows,0][:,None],n[rows,0][:,None])
        selected[rows]=pp.argmax(axis=1)+1
        if target!=400: # after current data before next allocation; mimic early looks at current N
            delta=pp_abs_reduction(y[rows,1:],n[rows,1:],y[rows,0][:,None],n[rows,0][:,None],delta=0.15)
            stop_fut=delta.max(axis=1)<early_futility
            stop_eff=pp.max(axis=1)>early_success
            if np.any(stop_fut):
                rr=rows[stop_fut]; active[rr]=False; fut[rr]=True
            if np.any(stop_eff):
                rr=rows[stop_eff]; active[rr]=False; success[rr]=True
        # allocate next block to rows still active
        rows=np.where(active)[0]
        if len(rows)==0: break
        current_n=n[rows].sum(axis=1)
        add_total=(target-current_n).astype(int)
        # if already at target? add_total valid
        # control add: target increments are 52/50/50/50, use 14 control except if add_total shorter
        cadd=np.minimum(n_ctrl_add,add_total)
        y[rows,0]+=rng.binomial(cadd,p[0]); n[rows,0]+=cadd
        aadd=add_total-cadd
        pp=pp_better(y[rows,1:],n[rows,1:],y[rows,0][:,None],n[rows,0][:,None])
        weights=pp/np.sqrt(n[rows,1:]+1)
        weights=np.where(weights<0,0,weights)
        weights=weights/weights.sum(axis=1,keepdims=True)
        weights=np.where(weights<0.05,0,weights)
        # if all zero, reset to equal
        row_sums=weights.sum(axis=1,keepdims=True)
        zero=(row_sums[:,0]==0)
        weights[~zero]=weights[~zero]/row_sums[~zero]
        weights[zero]=1/8
        # allocate counts for each distinct aadd (mostly 38/36)
        add_counts=np.zeros((len(rows),8),dtype=int)
        for val in np.unique(aadd):
            idx=np.where(aadd==val)[0]
            if val>0 and len(idx)>0:
                add_counts[idx]=allocate_counts(weights[idx],int(val))
        y[rows,1:]+=rng.binomial(add_counts,p[1:])
        n[rows,1:]+=add_counts
    # Final decision for any active rows at N=400
    rows=np.where(active)[0]
    if len(rows)>0:
        pp=pp_better(y[rows,1:],n[rows,1:],y[rows,0][:,None],n[rows,0][:,None])
        selected[rows]=pp.argmax(axis=1)+1
        success[rows]=pp.max(axis=1)>threshold
        active[rows]=False
    N=n.sum(axis=1).astype(float)
    top4=np.argsort(p[1:])[:4]+1
    pct_top4=n[:,top4].sum(axis=1)/N
    return eval_results(p,success,N,selected,pct_top4=pct_top4,futility=fut)

def calibrate(sim_func, target=0.10, nsim=12000, seed_base=11, **kwargs):
    lo,hi=0.80,0.9995
    for it in range(15):
        mid=(lo+hi)/2
```

```
        res=sim_func(SCENARIOS['Null'],nsim=nsim,threshold=mid,seed=seed_base+it,**kwargs)
        if res['Pr_success']>target:
            lo=mid
        else:
            hi=mid
    return (lo+hi)/2

if __name__=='__main__':
    """Run the CATS Monte Carlo comparator simulation.

    The SAP FACTS results for the selected Bayesian adaptive RAR + 2D NDLM
    design are entered as constants. Alternative simplified comparator designs
    are simulated with transparent beta-binomial screening approximations and
    calibrated under the global null to approximately 10% false-success.
    """
    def sim_rar_strict(p, nsim=1000, threshold=0.97, seed=1):
        # Independent-arm Bayesian RAR comparator with stricter early success threshold.
        return sim_bayes_ind_rar(p, nsim=nsim, threshold=threshold, early_success=0.9995, seed=seed)

    thr_fixed = calibrate(sim_fixed_final, nsim=2000, target=0.10)
    thr_mams = calibrate(sim_mams, nsim=1500, target=0.10, futility_pp=0.55)
    thr_parallel = calibrate(sim_parallel_two_stage, nsim=1500, target=0.10, keep_pp=0.50, futility_pp=0.45)
    thr_rar = calibrate(sim_rar_strict, nsim=1500, target=0.10)

    rows = []
    scen_order = ["Null","All15","Realistic","Realistic2","Backwards","Backwards2","Flat Drop","One Good One Bad","Harm10"]
    designs = [
        ("Fixed final-only Bayesian screen", sim_fixed_final, thr_fixed, {}),
        ("MAMS/drop-the-loser screen", sim_mams, thr_mams, {"futility_pp":0.55}),
        ("Parallel two-stage arm screens", sim_parallel_two_stage, thr_parallel, {"keep_pp":0.50,"futility_pp":0.45}),
        ("Bayesian independent-arm RAR screen", sim_rar_strict, thr_rar, {}),
    ]
    for scen in scen_order:
        p = SCENARIOS[scen]
        for name, func, thr, kwargs in designs:
            res = func(p, nsim=3000, threshold=thr, seed=100000 + int(hashlib.md5(f'{scen}|{name}'.encode()).hexdigest()[:8], 16) % 100000, **kwargs)
            rows.append({"Scenario":scen, "Design":name, "Threshold":thr, **res})
        rows.append({"Scenario":scen, "Design":"CATS Bayesian adaptive RAR + 2D NDLM (SAP FACTS)", "Threshold":np.nan, **SAP_FACTS[scen], "Pr_select_true_best":np.nan,
"Pct_top4_alloc":np.nan, "Pr_futility":np.nan})
        if scen in SAP_FIXED:
            rows.append({"Scenario":scen, "Design":"CATS fixed equal allocation (SAP)", "Threshold":np.nan, **SAP_FIXED[scen], "Pr_select_true_best":np.nan, "Pct_top4_alloc":np.nan,
"Pr_futility":np.nan})

    df = pd.DataFrame(rows)
    df.to_csv('/mnt/data/cats_monte_carlo_design_comparison.csv', index=False)
    print(df.to_string(index=False))
```
